# A panel of four miRNAs (miR-190b, miR-584-5p, miR-452-5p, and miR-1306-5p) is capable of classifying luminal and non-luminal breast cancers

**DOI:** 10.1101/2022.10.18.22281125

**Authors:** Faranak Farahmand, Saied Rahmani, Hadi Bayat, Adel Salimi, Sogol Ghanbari, Afsaneh Malekzadeh Shafaroudi, Ali Sharifi-Zarchi, Mohammad Vasei, Seyed-Javad Mowla

## Abstract

**BACKGROUND:** Identifying the molecular subtypes of breast cancer (BC) plays a crucial role in enhancing the efficacy of therapy. MiRNAs (miRs) with differential expressions in different subtypes of breast tumors can be considered as non-invasive biomarkers for diagnosing BC subtypes.

**OBJECTIVE:** We aimed to investigate the efficacy of miR-190b, miR-584-5p, miR-452-5p, and miR-1306-5p as novel potent diagnostic biomarkers in discriminating patients with luminal (ER+) and non-luminal (ER–) BCs.

**METHODS:** A group of miRs significantly associated with estrogen cell receptors (ER) in breast tumors were identified using feature selection methods analysis on miR-Seq datasets retrieved from TCGA and GSE68085. Four abovementioned miRs were selected as novel potential biomarkers, and their relative expression levels were assessed within adjacent non-tumor, ER+ and ER– tumor tissues by quantitative RT-PCR. Their impact on diagnosis was also evaluated by ROC curve analysis.

**RESULTS:** In ER+ BCs compared to ER– BCs, the expression of miR-190b was remarkably increased, while the expression of miR-584-5p, miR-452-5p, and miR-1306-5p were significantly decreased. This group could discriminate ER+ and ER– BCs at an AUC of 0.973.

**CONCLUSIONS:** According to our findings, these four miRs are promising biomarkers in discriminating BC subtypes. The candidate miRs in parallel with histologic diagnosis methods can be applied for identifying patients who are most likely responding to specific therapies based on ER status.

## 1 Introduction

Breast cancer (BC) has been the leading cause of cancer death (15.5%) and the most common cancer in females (24.5%) worldwide [1]. Despite significant improvements in the understanding of cancer pathogenesis, screening programs for early diagnosis, and treatment over the past few decades, there are still about 2.2 million new cases of BCs with more than half a million BC-related deaths recorded annually worldwide [1]. BC has heterogeneous nature with various morphologic signs and clinical outcomes which can be categorized in several aspects, including clinical features, expression of tumor markers, and histologic types [2, 3]. In this regard, gene expression profile analyses have led to the classification of BCs based on hormone receptors, including estrogen receptor (ER), progesterone receptor (PR), and human epidermal growth factor receptor (HER2 or HER2/neu) status. The hormone receptors are crucial factors in therapeutic prediction and should be measured on all newly diagnosed BC tumors [4-7]. ER and PR mediate mammary cell proliferation signals and stimulate the growth of both normal and neoplastic breasts [8]. Likewise, HER2 is a transmembrane receptor tyrosine kinase in the epidermal growth factor receptor family [8]. Accordingly, the intrinsic molecular subtypes of BC were classified as luminal A (ER+/PR±/HER2–), luminal B (ER+/PR±/HER2+), HER2 enriched (ER–/PR–/HER2+), basal-like, and normal-like, which show specific biological features and clinical outcomes [6].

In the clinical aspect, the ER expression is frequently examined by immunohistochemistry of biopsy to separate BC subtypes. This examination provides information on prognosis and the possibility of response to endocrine therapy [9]. However, the main disadvantage of this assay is false-negative results, which contribute to patients being denied for hormone therapy. Furthermore, diagnosis based on tissue biopsy is also an invasive process with the risk of spreading tumor cells to adjacent tissues [10, 11]. Therefore, considering the limitations of common diagnostic tests, developing new assays with high sensitivity and specificity for BC subtypes determination is indispensable. In this regard, biomarkers are now investigated in cancer studies for the purpose of diagnosis, prognosis, and therapy [12].

MiRNAs (miRs) are a class of small non-coding RNAs with 19–25 nucleotides length. The miRs can post-transcriptionally regulate gene expression by binding to the target mRNA and affecting many biological processes such as differentiation, proliferation, apoptosis, and metastasis [13]. Dysregulated miRs play primary roles in cancer initiation and progression [12, 13]. During recent years, an increasing number of miRs acting either as oncogenes or tumor suppressors have been investigated in BC [14]. Due to remarkable stability and easily non-invasive detection in biological fluids, such as serum and plasma, miRs are considered as promising biomarkers in cancer diagnosis and prognosis [12, 15-18]. Hence, numerous studies have reported expression patterns of miRs as an informative tool for the classification of BCs [19, 20]. Although several studies have been tried to discover biomarkers based on miRs, the exploration of novel groups of miRs with high sensitivity and specificity for the diagnosis of BC subtypes is valuable for developing diagnosis strategies, specific treatment, and disease management. The main purpose of the present study was to identify novel promising miR biomarkers which are associated with the presence of ER in BCs. For this aim, we utilized boruta [21], XGBoost [22], and limma [23] R packages for TCGA and GEO datasets analysis. Moreover, quantitative RT-PCR (qRT-PCR) method was used to validate experimentally the efficacy of our candidate miR biomarkers in discriminating luminal (ER+) from non-luminal (ER–) BCs. Furthermore, we evaluated the role of the candidate miRs as biomarker for BC diagnosis by measuring their expression in breast tumor and non-tumor samples of the TCGA dataset and those samples which were used in the qRT-PCR analysis. Finally, we confirmed the efficacy of miR-190b (miR-190b is classified as miR-190b-3p and -5p in the latest version of miRBase sequence database (release 22.1), Here, our candidate is miR-190b-5p), miR-584-5p, miR-452-5p, and miR-1306-5p as the promising biomarkers of luminal (ER+) and non-luminal (ER–) breast tumors.

## 2 Materials and methods

### 2.1. Analysis of miR-Seq datasets

Raw read counts of miR-seq dataset of BC patients along with their clinical dataset from TCGA [24] were obtained and analyzed by TCGAbiolinks package [25]. The total number of samples in this dataset was 1175 (1072 tumor and 103 adjacent non-tumor samples). Intrinsic subtype labels were assigned to samples according to the expression level of ER, PR, and HER2 from IHC test results reported in clinical data. Information about the BC subtypes based on the cell receptors (ER, PR, and HER2) was presented in positive/negative and level of existence format. In addition to TCGA, raw read counts of the miR-seq dataset of patients with TNBC and luminal (ER+) BCs were obtained from GSE68085 [26].

Here, we applied boruta feature selection, XGBoost feature selection, and limma differential expression analysis on TCGA and GSE68085 datasets. To merge the results of different feature selection methods on a dataset in a statistically acceptable manner, the Stuart method from the ‘RobustRankAggreg’ package in R was used. Also, in the other cases that we need to merge multiple ordered lists, we utilize the same strategy. After investigating miRs that are significantly associated with the existence of cell receptors in breast tumors, we used the ggplot2 R package to generate box plots by which we could evaluate the differential expressions of the top ten miRs with the highest association to ER between luminal (ER+) and non-luminal (ER–) breast tumor samples in miR-seq datasets. We also reviewed the related previous studies to identify those miRs which have not been previously reported as biomarkers for BC subtypes. A group of miRs which have not been previously reported as biomarkers for BC subtypes, were selected.

### 2.2. Patients and samples

This research involved using archived samples that were collected from October 2018 to June 2019 and the study were conducted from October 2020 to September 2021. BC tissue specimens were collected from Khatam-ol-Anbia and Rasule-Akram hospitals, they were immediately snap-frozen in liquid nitrogen after surgery and were stored at -80°C at Tarbiat Modares University. Tissue samples were categorized into 36 pairs of breast tumors and their adjacent non-tumor tissues, plus 13 breast tumor samples. All tumor samples were examined by pathologists and classified according to the standard histopathological parameters. Clinicopathological characteristics of patients are summarized in **Supplementary Table 1**. The research protocol for in vitro experiments on tissue samples was approved by the ethics committee of Ferdowsi University of Mashhad (code number: IR.UM.REC.1399.104). No experiments were conducted on human subjects or animals. Authors have no access to the information that could identify individual participants during or after data collection.

**Table 1.** Top-ranked most strongly ER associated miRNAs.

| ER | Aggregate Score |
| --- | --- |
| **miR-190b** | 0.000032 |
| miR-18a-5p | 0.000256 |
| **miR-505-3p** | 0.002304 |
| miR-224-5p | 0.00484 |
| **miR-577** | 0.005816 |
| miR-135b-5p | 0.007 |
| **miR-584-5p** | 0.01024 |
| **miR-452-5p** | 0.065408 |
| **miR-452-3p** | 0.075656 |
| **miR-1306-5p** | 0.114264 |

The results of the RobustRankAggreg package analysis of miRseq datasets are summarized in this table.
MiRNAs with a lower aggregate score are associated to ER receptors with a higher probability. Our
candidate miRNAs are in bold style.

### 2.3. RNA extraction

For isolating the total RNA from breast tissues, RiboEx Total RNA reagent (GeneAll Biotechnology, South Korea) was used. The concentration of the extracted RNA was then quantified using a NanoDrop™ spectrophotometer. The purity of the RNA was validated by measuring the ratio of the absorbance at 260 and 280 nm. The quality of RNA, which is regarded as the absence of degraded RNA, was evaluated by agarose gel electrophoresis and ethidium bromide staining. Accordingly, the 18S and 28S RNA bands were visualized under ultraviolet light.

### 2.4. Polyadenylation and cDNA synthesis

Following RNA isolation, 1 μg of total RNA was poly-adenylated using Poly(A) Polymerase Tailing Kit (New England Biolabs., UK., Ltd.) according to manufacture protocol. Then, the poly-adenylated RNA (10 μl) was converted to complementary DNA (cDNA) by adding Anchored Oligo(dT) and using RevertAid M-MuLV RT (Thermo Fisher Scientific., UK) as the protocol provided by corresponding manufacture. The prepared cDNA was used for quantification of miR-190b, miR-584, and miR-1306.

### 2.5. Stem–loop RT–PCR and cDNA synthesis

Stem-loop reverse transcription was performed for miR-452 using stem-loop primers. Then, reverse transcriptase reaction was performed using RevertAid M-MuLV RT (Thermo Fisher Scientific., UK).

### 2.6. qRT-PCR

Ultimately, qRT-PCR was performed to quantitatively assess the expression of our selected miRs in tissue samples using appropriate primer sets (**Supplementary Tables 2 and 3)**. Syber Green PCR Master Mix (BIOFACT Co., Ltd., Korea) and primers listed in **(Supplementary Tables 2 and 3)** were utilized for qRT-PCR, which was conducted on a StepOne Plus System. Mean delta Ct values of triplicate real-time qRT-PCR amplifications were utilized in statistical analysis. The comparative delta Ct values (deltaCt (ΔCt) = Ct_miRs_ - Ct_U-48_) and Log 2^-^ ΔCt were used as the relative quantification of miRNAs, using the U48 small RNA [27-29] for normalization. All the amplified candidate miRs which TA-cloned using pGEM-T easy vector kit (Promega; USA). Finally the accuracy of cloned sequences were confirmed by the golden standard sequencing method.

### 2.7 Statistical analysis

GraphPad Prism ver. 8 (GraphPad Software Inc., La Jolla, CA, USA) was applied for statistical analysis of the results obtained by qRT-PCR. The two-tailed Mann-Whitney test was used to compare the differential expression level of selected miRs within the ER+ and ER– breast tumor samples.

The Wilcoxon test was used for comparison of our selected miRs expression level between breast tumor and their adjacent non-tumor tissues. Results with p-value < 0.05 were considered as significant.

### 2.8 ROC Curve analysis

Receiver operating characteristic (ROC) curves were also plotted by the pROC package [30] to validate the capability of the candidate miRs to distinguish between ER+ and ER– tumor samples and between breast tumors and their adjacent non-tumor samples. This was performed both individually and for a combination of all selected miRs. The area under curve (AUC) was also employed for evaluation of the specificity and sensitivity of the candidate miRs in distinguishing ER+ and ER– tumor samples and their adjacent non-tumor tissues; the higher AUC shows better diagnostic performance (the AUCs closer to 1 reflect more substantial differences).

## 3 Results

### 3.1. The candidate miRs associated with ER in breast tumors

The miRs with the highest possibility of being associated with ER were identified and ordered using limma, xgboost, bruta and aggregated by the RobustRankAggreg package. Boruta is an R package utilizing a random forest model to classify data. XGBoost is also an R package that provides a regression and classification model based on tree models and the ensemble technique. Further, we combined all the results to reach the most generalizable miR biomarkers. We suppose that several miRs are significantly associated with the existence of cell receptors; therefore, this association should be found in most of the datasets and by the majority of methods without considering methods and experiment biases. The final result of a feature selection method on a dataset is an ordered list of miRs. As we aimed to investigate potential biomarkers of luminal (ER+) and non-luminal (ER–) breast tumors, we focused our study on the top ten most strongly ER associated miRs **(Table 1)**. The expression status of these top ten miRNAs in BC were investigated in UCSC genome browser (GRCH37/hg19), and their differential expression were investigated in ER+ and ER– breast tumor samples of TCGA **(Supplementary Fig. 1)**. We excluded miR-577 and miR-452-3p from the top ten list due to their extremely low expression levels in breast tumors. The first rank miR-190b with the highest score in the list was selected as the potential biomarker of ER+ breast tumors. The expression levels and the association of miR-18a-5p [31], miR-505-3p [32], miR-224-5p [33, 34], and miR-135b-5p [35] with ER have been previously investigated in breast cancer. Therefore, we excluded miR-18a-5p, miR-505-3p, miR-224-5p, and miR-135b-5p from this study, and we selected those miRs which their association with highly aggressive breast tumors (ER–) have not been previously studied. In this regard, miR-584-5p, miR-452-5p, and miR-1306-5p were selected to evaluate their potential as biomarkers of ER– breast tumors.

### 3.2. Evaluation of miR-190b expression level in ER+ compared to ER– breast tumors

Our *in-silico* analysis on the miR-seq datasets in TCGA **(Fig. 1A)** and GSE68085 **(Fig. 1B)** databases, elucidated higher expression level of miR-190b in ER+ compared to ER– breast tumors (P. Value < 0.05). Moreover, as shown in **Fig. 1C**, the expression level of miR-190b correlated positively with the percentage of ER level in breast tumors, which indicates that the higher the ER level is, the more miR-190b is expressed in breast tumors. The results of qRT-PCR confirmed our *in-silico* analysis **(Fig. 1D)**. Indeed, we found a similar significant higher expression level of miR-190b in 24 ER+ compared to 23 ER– breast tumor samples (P. Value < 0.05).

**Fig. 1:**
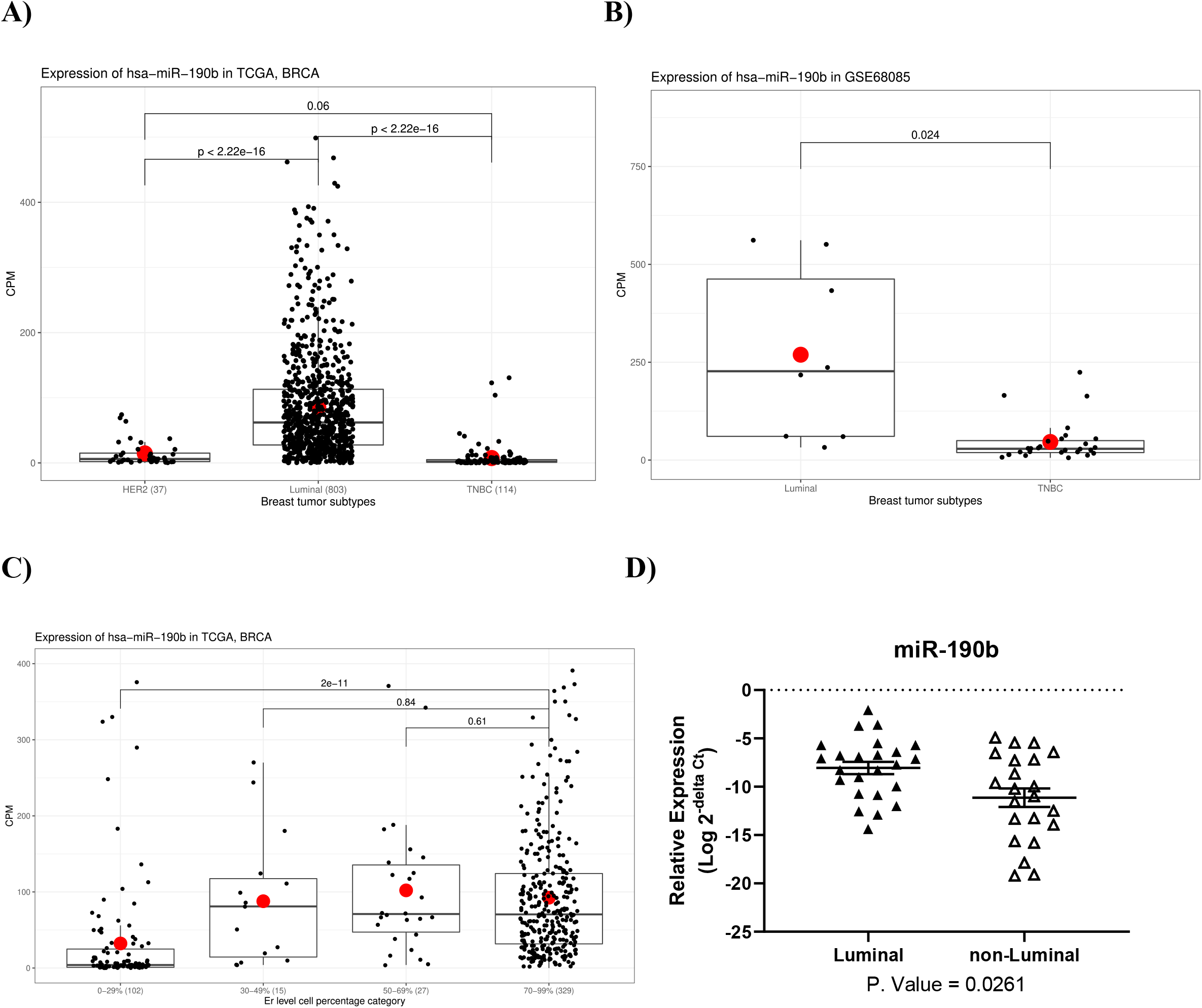
The expression of miR-190b in patients with ER+ compared to ER– BCs. The *in-silico* analysis of the miR-seq data of BC patients in TCGA **(A)** and GSE68085 **(B)** showed a significant upregulation of miR-190b in ER+ versus ER– breast tumor samples (P. Value < 0.05). **C)** The *in-silico* evaluation of miR-190b expression based on ER percentage level showed a positive correlation between miR-190b and ER expression level. **D)** The results of the qRT-PCR analysis also showed that the expression of miR-190b (relative to u-48) in ER+ breast samples is significantly higher than ER– breast samples (P. Value < 0.05).

### 3.3. Evaluation of the expression of miR-584, miR-452, and miR-1306 in ER+ compared to ER– breast tumors

We validated the significant downregulation of miR-584, miR-452, and miR-1306 in ER+ compared to ER– breast tumors by qRT-PCR **(Fig. 2D)**, which was predicted by analyzing the available miR-seq datasets in TCGA **(Fig. 2A)** and GSE68085 **(Fig. 2B)** (P. Value < 0.05). In *in-silico* analysis, a negative correlation between the percentage of ER level and the expression of miR-584, miR-452, and miR-1306 was also observed in breast tumors **(Fig. 2C)**.

**Fig. 2:**
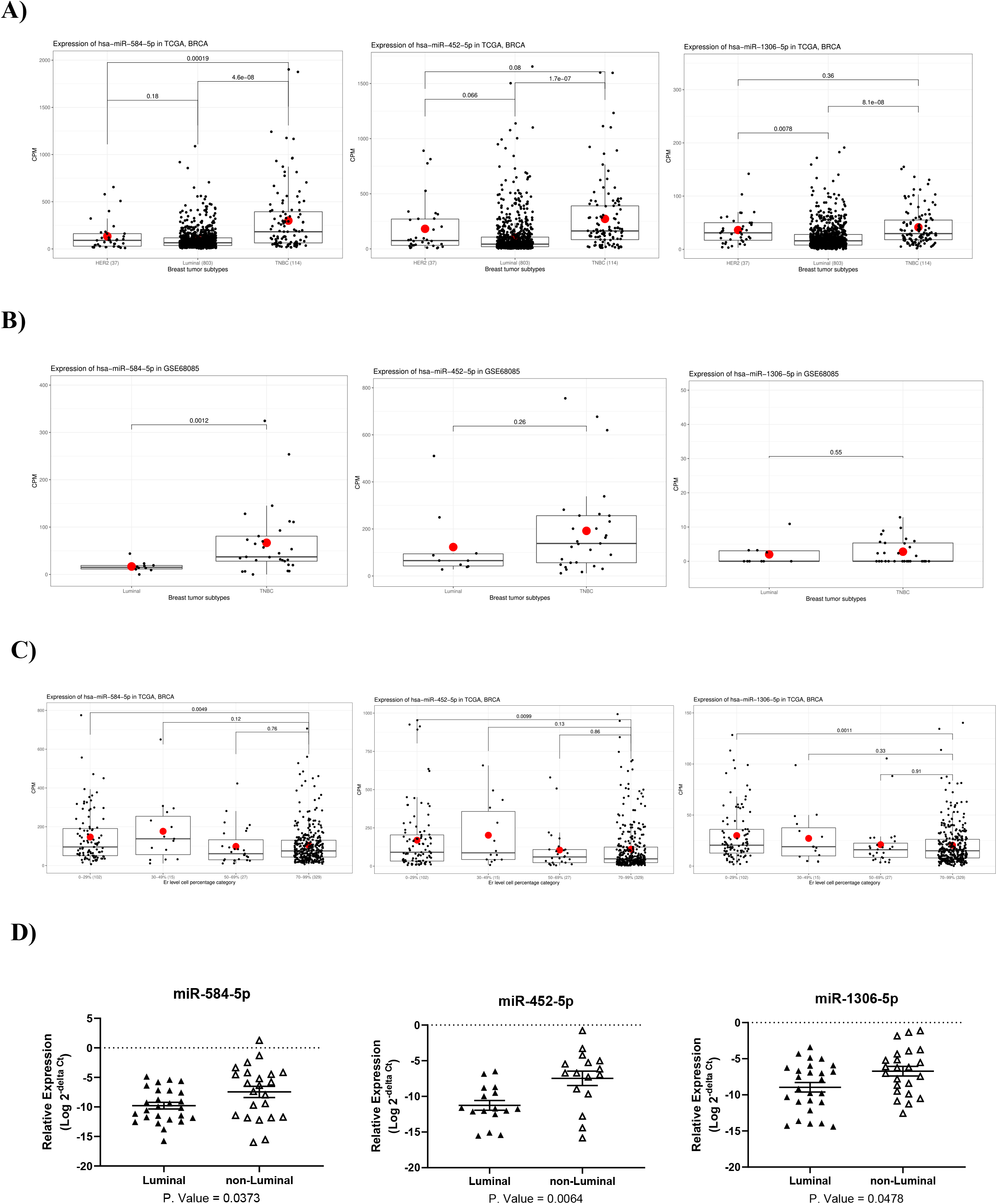
The expression of miR-584, miR-452, and miR-1306 in patients with ER+ compared to ER– BCs. The results of *in-silico* analysis of the miR-seq dataset of BC patients in TCGA **(A)** and GSE68085 **(B)** showed downregulation of miR-584, miR-452, and miR-1306 (P. Value < 0.05) in ER+ versus ER– breast tumor samples. **C)** The *in-silico* evaluation of the expression of miR-584, miR-452, and miR-1306 based on the percentage of ER level indicated a significant negative correlation between these three miRs and ER expression level (P. Value < 0.05). **D)** The qRT-PCR analysis also showed that the expression of miR-584, miR-452, and miR-1306 (relative to u-48) in ER+ breast samples is significantly lower than ER– breast samples (P. Value < 0.05).

### 3.4. Evaluation of Differential expression of our four selected miRs between breast tumor and non-tumor samples

At first, expression profiles of the selected miRs between breast tumor and non-tumor samples (adjacent non-tumor breast tissues) in TCGA datasets were determined by *in-silico* analysis. Then, the expression of the candidate miRs were evaluated experimentally in 36 breast tumor cases and their adjacent non-tumor tissues by qRT-PCR analysis. Although *in-silico* evaluation of miR-190b expression in breast tumors compared to non-tumor breast samples of TCGA data showed a significant upregulation **(Fig. 3A)**, the qRT-PCR analysis showed an opposite results **(Fig. 3B)**. Further analysis indicated that the downregulation of miR-190b in breast tumors was restricted to ER−/PR− tumors while ER+/PR+ and ER+/PR− tumors showed the upregulation of miR-190b compared to non-tumor breast tissues **(Fig. 3C and D)**. Therefore, the upregulation or downregulation of miR-190b in tumor samples versus non-tumor samples is dependent on the ER status.

**Fig. 3:**
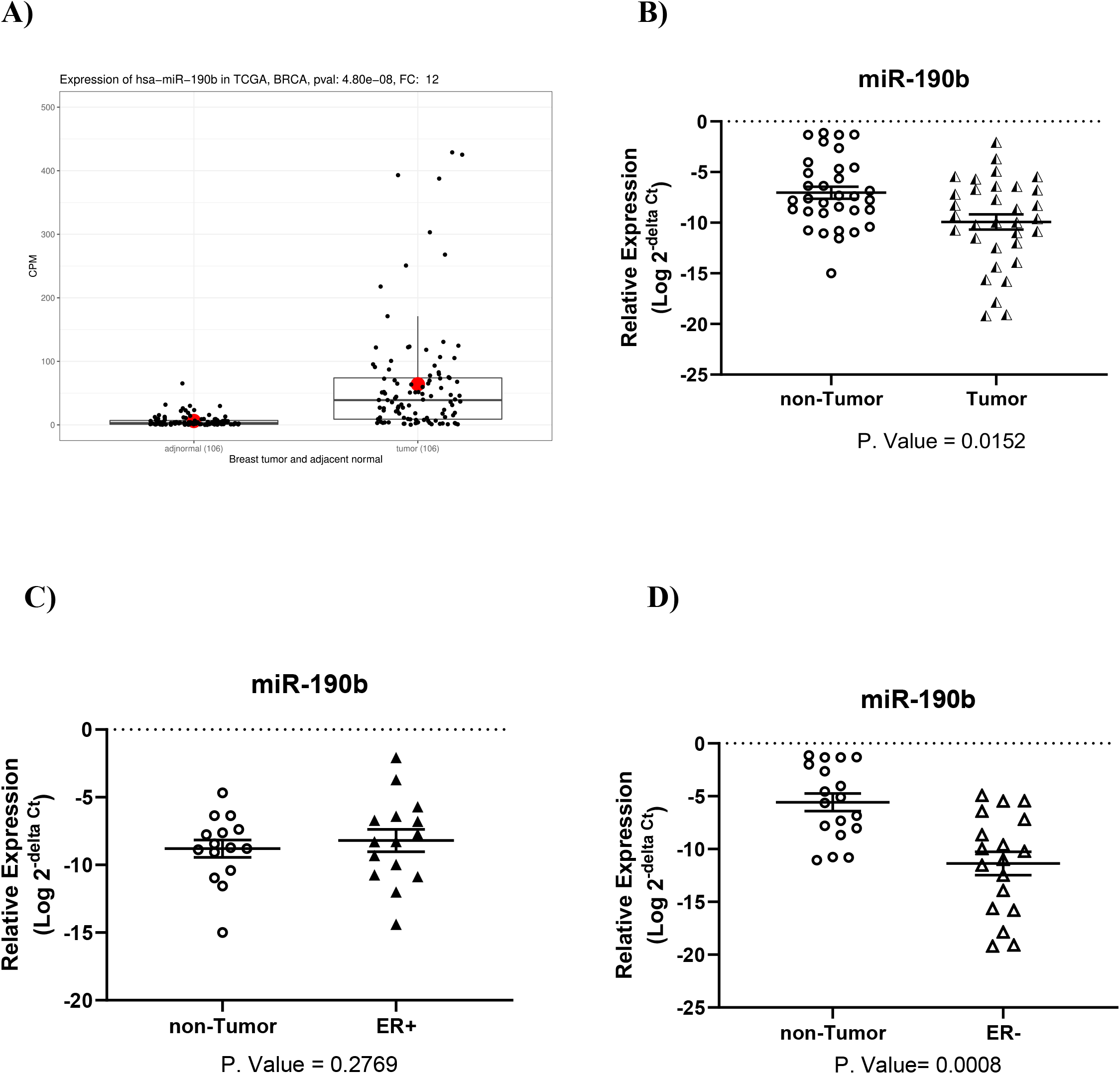
The expression of miR-190b (relative to U-48) in breast tumor samples compared to non-tumor adjacent tissues. **A)** The results of *in-silico* analysis showed a significant increase of miR-190b expression in breast tumors compared to non-tumor breast samples of TCGA data (P. Value < 0.05). **B)** However, the results of the qRT-PCR analysis showed an overall decrease of miR-190b expression in tumor tissues versus non-tumor adjacent tissues (P. Value < 0.05). The qRT-PCR analysis also showed that miR-190b was up-regulated in ER+ tumors **(C)**, whereas it was downregulated in ER– tumors compared to non-tumor adjacent tissues **(D)**.

MiR-584-5p, miR-452-5p, miR-1306 showed lower expression in breast tumor samples (both ER+ and ER–) compared to non-tumor breast tissues based on both miR-seq **(Fig. 4A, B, and C)** and qRT-PCR analysis **(Fig 4D, E, and F)**. Although, in *in-silico* analysis of miR-seq datasets, miR-1306 didn’t show significant differential expression between breast tumor and non-tumor adjacent samples **(Fig. 4C)**, in the qRT-PCR analysis, it was significantly down-regulated in breast tumor tissues compared to their adjacent non-tumor tissues (p-value < 0.05) **(Fig. 4F)**.

**Fig. 4:**
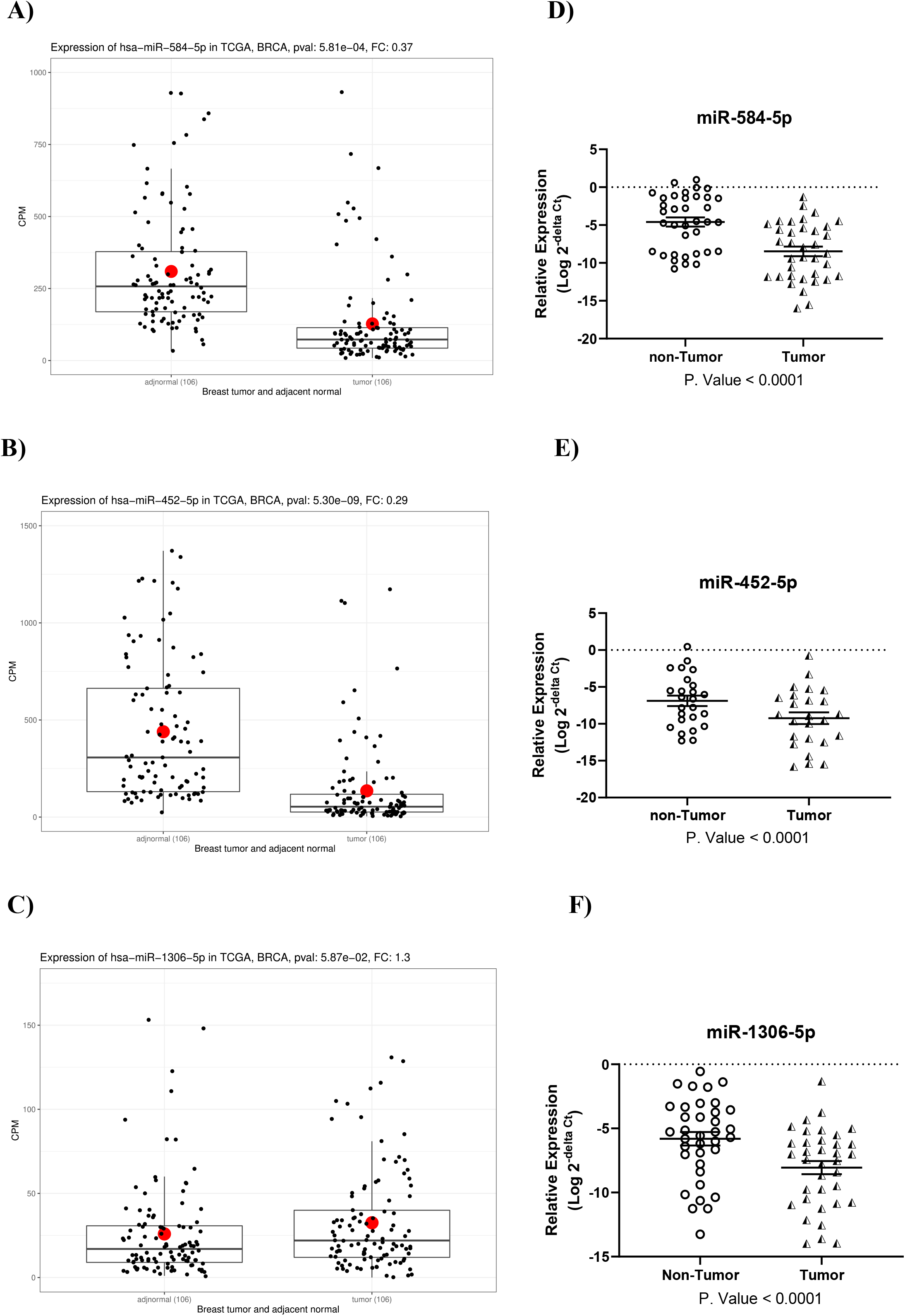
The expression of miR-584, miR-452, and miR-1306 (relative to U-48) in breast tumor samples compared to non-tumor adjacent tissues. The results of in-silico analysis showed that the expression of miR-584 **(A)**, and miR-452 **(B)** are significantly lower in breast tumors, as compared with non-tumor breast samples of TCGA (P. Value < 0.05), while the expression level of miR-1306 **(C)** showed no significant difference between breast tumors and non-tumor breast samples. The results of the qRT-PCR analysis showed significant downregulation of miR-584 **(D)**, miR-452 **(E)**, and miR-1306 **(F)** in tumor tissues versus non-tumor adjacent tissues (P. Value < 0.05).

### 3.5. Roc Curve analysis

The four candidate miRs, miR-190b, miR-584, miR-452 and miR-1306, showed high AUCs with values of 0.951 (specificity of 86%, and sensitivity of 94%), 0.851 (specificity of 82%, and sensitivity of 75%), 0.846 (specificity of 79%, and sensitivity of 81%), and 0.80 (specificity of 72%, and sensitivity of 78%), respectively **(Fig. 5A, B, C, and D)**. Interestingly, the AUC of the combination of miR-190b, miR-584, miR-452, and miR-1306 increased to 0.973 with the specificity of 92% and sensitivity of 96% in discriminating ER+ and ER– samples of TCGA dataset **(Fig. 5E)**. A ROC curve was also plotted for the combination of miR-190b, miR-18a-5p, miR-505-3p, miR-224-5p, miR-135b-5p, miR-584-5p, miR-452-5p, and miR-1306-5p which created the similar AUC value of 0.977 **(Supplementary Fig. 2)**. Therefore, the addition of miR-18a-5p, miR-505-3p, miR-224-5p, and miR-135b-5p did not add values to the AUC achieved by the combination of our selected miRs (Both groups represented the AUC of 0.97). In addition to the ROC curves obtained by the in silico analysis of TCGA datasets, the pROC package was used to evaluate the diagnostic value of our selected miRs in discriminating ER+ from ER– tissue samples used in the qRT-PCR analysis. In concordance with AUCs obtained for miR levels in TCGA datasets, significant AUCs of 0.961 (Specificity of 71%, and sensitivity of 63%), 0.674 (Specificity of 69%, and sensitivity of 61%), 0.777 (Specificity of 87%, and sensitivity of 69%), and 0.666 (Specificity of 77%, and sensitivity of 52%) have been obtained for miR-190b **(Fig. 5F)**, miR-584 **(Fig. 5G)**, miR-452 **(Fig. 5H)**, and miR-1306 **(Fig. 5I)** respectively (P. Value < 0.05).

**Fig. 5:**
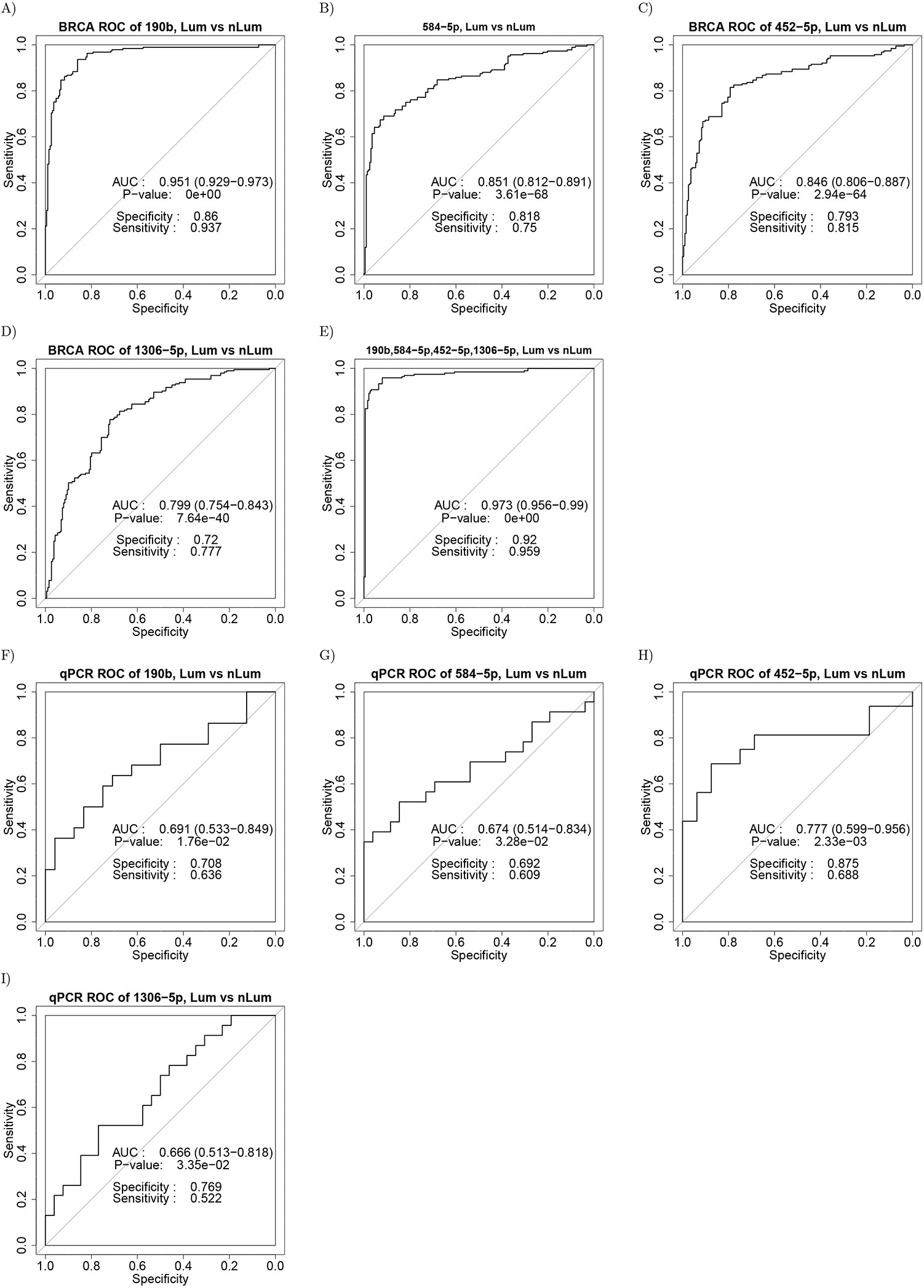
The AUCs of ROC curves for miR levels in ER+ and ER– samples. The capabilities of miR-190b **(A** and **F)**, miR-584 **(B** and **G)**, miR-452 **(C** and **H)**, miR-1306 **(D** and **I)**, and the combination of these four miRs **(E)** to discriminate ER+ and ER– samples of TCGA datasets **(A, B, C, D**, and **E)** and qRT-PCR analysis **(F, G, H**, and **I)** are shown in ROC curves.

As shown in the qRT-PCR results, miR-190b was up-regulated in ER+ breast tumors, whereas it was down-regulated in ER– tumors compared to non-tumor tissues. Therefore, we excluded this miR from the group of biomarkers which efficiency was evaluated in the detection of breast tumors. Accordingly, ROC curves were plotted for miR-584, miR-452, and miR-1306 expression in TCGA datasets, which represented the AUCs of 0.961 (specificity of 90%, and sensitivity of 91%), 0.926 (specificity of 93%, and sensitivity of 82%), and 0.725 (specificity of 63%, and sensitivity of 74%) (**Fig. 6A, B, and C**). The AUCs of 0.939 (specificity of 97%, and sensitivity of 86%), and 0.982 (specificity of 91%, and sensitivity of 95%) were also generated for the combination of miR-584 and miR-452, and the combination of miR-584, miR-452, and miR-1306 (**Fig. 6D and E**).

**Fig. 6:**
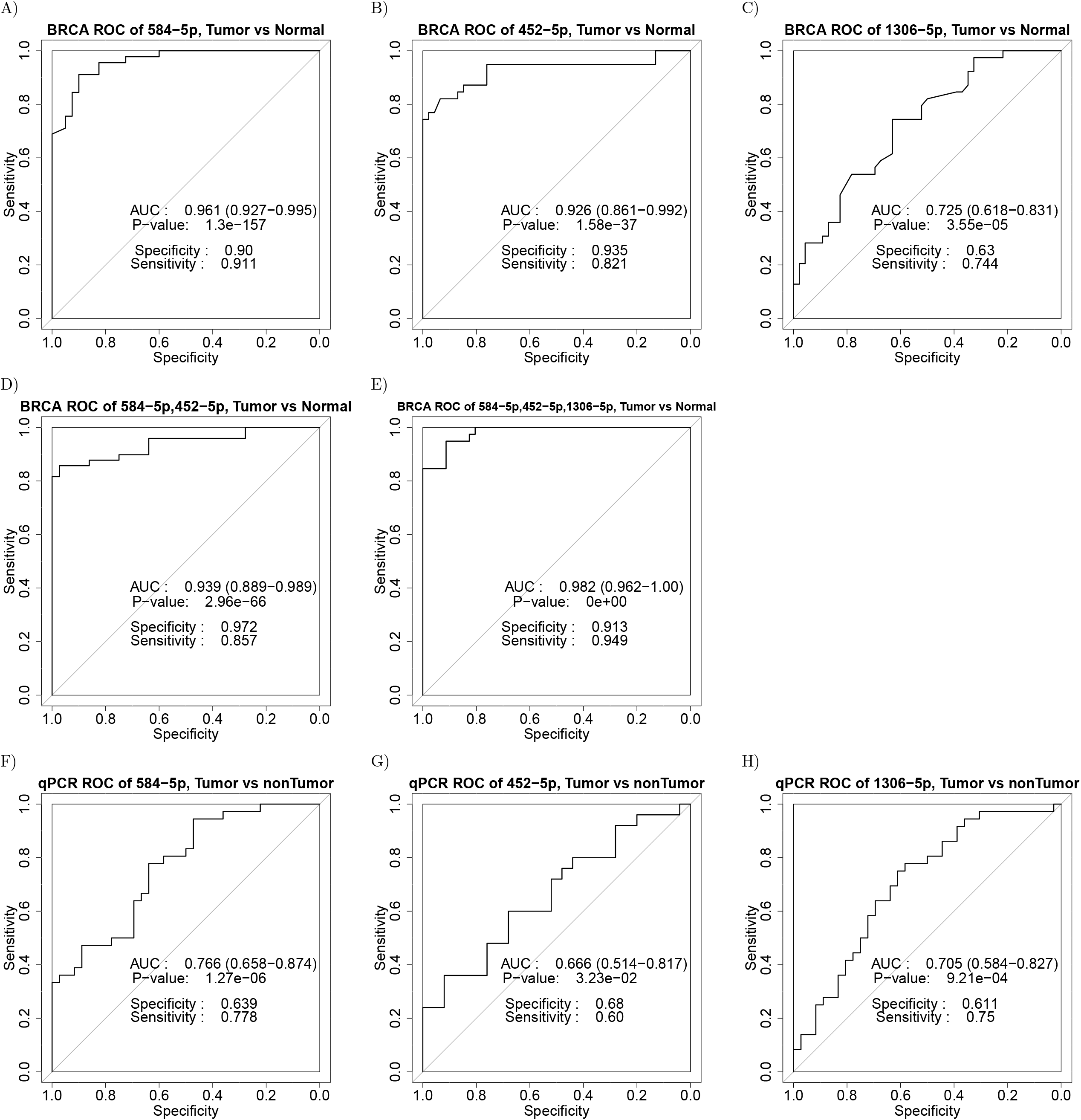
The AUCs of ROC curves for mi levels in tumor and non-tumor samples. The potential of miR-584 **(A** and **F)**, miR-452 **(B** and **G)**, miR-1306 **(C** and **H)**, the combination of miR-584 and miR-452 **(D)**, and the combination of all these three miRs **(E)** to discriminate tumor and non-tumor samples of TCGA **(A, B, C, D, E)** and qRT-PCR analysis **(F, G, H)** are shown in ROC curves.

Furthermore, the ROC curves generated for the expression of miR-584, miR-452, and miR-1306 in tumor and adjacent non-tumor samples of qRT-PCR analysis represented the AUC values of 0.766 (specificity of 64%, and sensitivity of 78%), 0.666 (specificity of 68%, and sensitivity of 60%), and 0.705 (specificity of 61%, and sensitivity of 75%) **(Fig. 6F, G, and H)**.

## 4 Discussion

In the current study, we aimed to identify ER associated miRs to reach promising biomarkers. Therefore, we used the available data from TCGA and applied boruta and XGBoost feature selection and limma differentially expression analysis to obtain miRs that are significantly associated with the existence of cell receptors in breast tumors. As a result, we reported the discovery of four miRNAs, miR-190b, miR-584-5p, miR-452-5p, and miR-1306-5p, which are significantly associated with ER status in breast tumors, as predicted by *in silico* analysis. We then confirmed the efficacy of this signature in discriminating luminal BCs (ER+) and non-luminal BCs (ER–) by qRT-PCR.

Currently, Mammography is the golden standard tool for screening and diagnosis of breast cancer (BC). However, invasive histological evaluation of breast biopsy is required for accurate diagnosis of the BC subtype. The identification of novel, reliable, and minimally invasive BC biomarkers with high sensitivity and specificity would lead to a significant improvement in the clinical management of this complex disease [36]. Numerous studies have reported the significant role of miRs in the initiation and progression of BC and revealed that certain miRs are differentially expressed between different breast tumor subtypes. [37]. Hence, the differentially expressed miRs can be considered as a promising molecular biomarkers for distinguishing BC subtypes.

It has been reported that miR-190b is associated with ER+ breast tumors [38] and resistance to hormone therapy [39]. Furthermore, Cizeron-Clairac et al. reported that miR-190b is significantly upregulated in ER+ compared to ER– tumors. In consistent with previous studies, our results also demonstrated the significant higher expression level of miR-190b in ER+ compared to ER– breast tumors (P. Value < 0.05). Moreover, ROC curve analysis showed the potential diagnostic value of miR-190b as a very remarkable biomarker in distinguishing ER+ from ER– breast tumors with the AUC of 0.951 (based on *in-silico* analysis). In another study, the expression of miR-190b was examined in seven BC cell lines and suggested that the biological and clinical implication of miR-190b may differ among BC subtypes [40]. Furthermore, de Anda-Jáuregui et al. showed the upregulation of miR-190b in ER+ breast tumors compared to normal breast tissues, whereas the downregulation of miR-190b in ER– tumors compared to healthy controls [38]. In the present study, our *in-silico* analysis represented an overall upregulation of miR-190b in breast tumors versus adjacent non-tumor tissue. While in the practical experiment, we observed the upregulation of miR-190b in ER+ but not in ER– breast tumors compared to non-tumor tissues. Hence, it should be noted that the upregulation of miR-190b in breast tumor samples versus non-tumor adjacent tissues was restricted to ER+ tumors. This discrepancy between the results of *in-silico* and qRT-PCR analysis may be attributed to the higher number of ER+ tumors compared to ER– tumors in the TCGA dataset. Previous studies mostly have focused on the role of miR-584 in lung cancer progression and metastasis [41-43]. The chromosomal region where miR-584 is located, 5q32, has been highlighted to be deleted in myelodysplastic syndromes that lead to malignant transformation [44, 45]. In addition, a study indicated that miR-584 may act as a tumor suppressor in renal carcinoma cells [46]. Furthermore, it was revealed that miR-584 is significantly down-regulated in human HER2+ breast tumors compared to non-tumor adjacent tissues [47]. Our results also showed significant downregulation of miR-584 in breast tumors compared to adjacent non-tumor tissues, which may indicate the tumor suppressive role of miR-584. According to ROC curve analysis, miR-584 also can act as a strong biomarker for BC diagnosis with high sensitivity and specificity (AUC of 0.961 and 0.766 in *in-silico* and qRT-PCR analysis, respectively). Additionally, our findings validated the downregulation of miR-584 in ER+ versus ER– breast tumors predicted by *in silico* analysis, and the AUC values of 0.851 and 0.674 obtained by *in-silico* and qRT-PCR analysis that represented the notable capability of miR-584 in distinguishing ER– and ER+ BC subtypes.

It is reported that miR-452 is expressed aberrantly in different types of human cancer [48-50]. The downregulation of miR-452 in BC tissues compared with paired normal breast tissues was also identified previously [51]. Moreover, miR-452 has been predicted to have an important role in regulation of pathways specific to luminal-A by TCGA data analysis [52]. In the present study, we also demonstrated the downregulation of miR-452 in breast tumors versus adjacent non-tumor tissues. More importantly, for the first time to our knowledge, we showed that miR-452 could serve as a potential biomarker for distinguishing ER– from ER+ breast tumors with high sensitivity and specificity (AUC values of 0.846 and 0.777 by *in-silico* and qRT-PCR analysis, respectively).

The aberrant expression of miR-1306 was observed in plasma samples of patients with different diseases such as heart failure, glaucoma, or epilepsy [53]. Moreover, the dysregulation of miR-1306 has been reported in human breast and colorectal cancers [53, 54]. Likewise, the upregulation of miR-1306-5p was observed in subjects with malignant breast lesions compared to benign tumors [53]. Our study represented the upregulation of miR-1306 in ER– breast tumors which typically are more aggressive than ER+ tumors. However, the downregulation of miR-1306 in breast tumors compared to non-tumor adjacent tissues may suggest its tumor suppressive role. This discrepancy needs to be elucidated in future studies. Likewise, no previous study has been performed to investigate the expression level and biological roles of miR-1306 in BC subtypes. Hence, this is the first study to our knowledge, that reveals the efficacy of miR-1306-5p in distinguishing ER+ and ER– breast tumors with AUC values of 0.799 and 0.666 based on *in-silico* and qRT-PCR analysis, respectively.

Overall, the AUCs acquired by qRT-PCR were not as high as those of *in-silico* analysis. This discrepancy can be attributed to the much higher number of breast tumor samples of TCGA datasets compared to the samples used in qRT-PCR. For identifying the best biomarker panel, we compared the AUCs produced from ROC curve analysis for each individual miR and their combination profiles in distinguishing ER+ and ER– BC samples of TCGA. It was revealed that the best AUC of 0.973 can be achieved from the combination of miR-190b, miR-584, miR-452, and miR-1306, providing specificity of 92% and sensitivity of 96%.

The ROC curve for the combination of the miR-584-5p, miR-452, and miR-1306 in breast tumors and non-tumor samples of TCGA showed an exceptionally high diagnostic accuracy with an AUC value of 0.982 with the specificity of 91% and sensitivity of 95%. It should be noted that miR-190b was not included in this biomarker set because its differential expression in breast tumors compared to non-tumor tissues depends on the tumor subtype. Although the signature of miR-584, miR-452, and miR-1306 is an effective diagnostic test alone with a strong performance (specificity of 100% and sensitivity of 87%), it may show promise as a valuable assessment tool in BC diagnosis in combination with screening mammography.

A particular strength of this study is that the reported mi panel is probably less prone to biological differences than a single miR. Therefore, the miR panels are more reliable for clinical use. A second strength is that the measured expression level of the four candidate miRs in breast tumor samples revealed a specific algorithm that helps to detect the ER status of each sample. To clarify, in ER+ tumors, the expression level of miR-190b was higher than that of miR-584, miR-452, and miR-1306, while, in ER– tumors the results were vice versa **(Supplementary Fig. 3)**. However, since the miR-452 expression level was not detectable in a number of tumor samples, the role of miR-452 in this algorithm needs further investigations.

## 5 Conclusion

Overall, our study signified the discovery of a panel of four miRs (miR-190b, miR-584-5p, miR-452-5p, and miR-1306-5p), which expression have association with ER in breast tumors, for diagnosis of luminal (ER+) and non-luminal (ER–) BC subtypes. According to this miR panel, we have represented a classification model with high discrimination ability in classifying ER+ and ER– BCs at an AUC of 0.973. Interestingly, a straightforward classification was introduced based on the seesaw expression pattern of miR-190b compared to the other three miRs to distinguish ER– and ER+ samples. In addition, the panel containing miR-584, miR-452, and miR-1306 was shown to have the efficacy to be developed as a parallel test to examine breast samples of patients with abnormal screening mammograms, with the aim of reducing false-positive results. For future investigations, these two miR signatures should be evaluated in blood serum samples to confirm the clinical utility of these miR signatures.

## Data Availability

All data produced in the present study are available upon reasonable request to the authors.

## Acknowledgments

Not applicable.

## Author contributions

**Conception:** Faranak Farahmand

**Interpretation or analysis of data:** Faranak Farahmand, Saeid Rahmani, Hadi Bayat

**Preparation of the manuscript:** Faranak Farahmand, Saeid Rahmani

**Revision for important intellectual content:** Faranak Farahmand, Saeid Rahmani, Seyed-Javad Mowla, Hadi Bayat, Adel Salimi, Sogol Ghanbari, Ali Sharifi-Zarchi, Mohammad Vasei

**Supervision:** Seyed-Javad Mowla

## Supporting information captions

**Supplementary Fig. 1:** The differential expression of miR-190b **(A)**, miR-18a-5p **(B)**, miR-505-3p **(C)**, miR-224-5p **(D)**, miR-577 **(E)**, miR-135b-5p **(F)**, miR-584-5p **(G)**, miR-452-5p **(H)**, miR-452-3p **(I)**, and miR-1306-5p **(J)** in ER+ versus ER– breast tumor samples of TCGA.

**Supplementary Fig. 2:** A ROC curve for the combination of miR-190b, miR-18a-5p, miR-505-3p, miR-224-5p, miR-135b-5p, miR-584-5p, miR-452-5p, and miR-1306-5p in ER+ and ER– breast tumor samples of TCGA which created the AUC value of 0.977.

**Supplementary Fig. 3:** The expression of miR-190b, miR-584-5p, miR-452-5p, and miR-1306-5p (relative to u-48) in ER–/PR– and ER+/PR+ breast tumor samples. The results of the qRT-PCR analysis revealed that the measured expression level of miR-190b is lower in each 9 ER– /PR–samples **(A)** and is higher in each 8 ER+/PR+ samples **(B)**, in comparison with the expression levels of the other three miRs in those samples (The expression of miR-452-5p was not detectable in T1, T2, T4, T8, T11, T12, T13, T14, T15, T16, and T17 samples).

## Notes

### Competing Interest Statement

The authors have declared no competing interest.

### Funding Statement

This study did not receive any funding.

### Author Declarations

The research protocol for in vitro experiments on tissue samples was approved by the ethics committee of Ferdowsi University of Mashhad (code number: IR.UM.REC.1399.104).

### Summary of Updates

The method section was updated.

