## Supplementary Table 1 for "A panel of four miRNAs (miR-190b, miR-584-5p, miR-452-5p, and miR-1306-5p) is capable of classifying luminal and non-luminal breast cancers"

**Supplementary Table 1.** Clinicopathological characteristics of patients

| **Clinicopathological characteristics** | | **Number of tumor samples** |
|---|---|---|
| **Subtype** | Luminal (ER+/PR±/HER2±) | 26 |
| | HER2+ | 15 |
| | TNBC | 8 |
| **Age** | <55 (31-54) | 22 |
| | ≥55 (55-75) | 27 |
| **Grade** | 0 | 1 |
| | 1 | 2 |
| | 2 | 25 |
| | 3 | 21 |
| | 4 | 0 |
| **Stage** | 0 | 1 |
| | 1 | 9 |
| | 2 | 20 |
| | 3 | 17 |
| | 4 | 2 |
