## Supplementary Table 2 for "A panel of four miRNAs (miR-190b, miR-584-5p, miR-452-5p, and miR-1306-5p) is capable of classifying luminal and non-luminal breast cancers"

**Supplementary Table 2.** List of primers for miR-190b, miR-584-5p, and miR-1306-5p

| Gene | Accession number | Primer | Sequence |
|---|---|---|---|
| miR-190b | MIMAT0004929 | Forward | 5’-GGCGGTGATATGTTTGATATTGG-3’ |
| miR-584-5p | MIMAT0003249 | Forward | 5’-CAGCGATTATGGTTTGCCTGG-3’ |
| miR-1306-5p | MIMAT0022726 | Forward | 5’-TCCACCTCCCCTGCAAAC-3’ |
| U-48 | NR_002745 | Forward | 5’-TGACCCCAGGTAACTCTGAGTGTGT-3’ |
| | | Universal Revers Primer | 5’-CCAGTGAGCAGAGTGACG-3’ |
| | | Anchored Oligo dt mix | 5’-GCGTCGACTAGTACAACTCAAGGTTCTTCCAGTCACGACG TTTTTTTTTTTTTTTTTT-3’ |

The comparative threshold cycle (Ct) was determined for each miRNA and the relative amount of each miRNA in individual samples was described as ΔCt (Ct miRNA- Ct internal control). $2^{-\Delta Ct}$ values were employed for expression level comparison of miRNAs in breast samples.
