## Supplementary Table 3 for "A panel of four miRNAs (miR-190b, miR-584-5p, miR-452-5p, and miR-1306-5p) is capable of classifying luminal and non-luminal breast cancers"

**Supplementary Table 3.** List of primers for miR-452-5p

| Gene | Accession number | Primer | Sequence |
|---|---|---|---|
| miR-452-5p | MIMAT0001635 | Forward | 5'-GGCGAACTGTTTGCAGAGG-3' |
| | | Stem-loop | 5'-GTCGTATCCAGTGCAGGGTCCGAGGTATTCGCACTGGATACGACTCAGTT-3' |
| U-48 | NR_002745 | Forward | 5'-TGACCCCAGGTAACTCTGAGTGTGT-3' |
| | | Stem-loop | 5'-GTCGTATCCAGTGCAGGGTCCGAGGTATTCGCACTGGATACGACGGTCAG-3' |
| | | Reverse | 5'- GTC GTA TCC AGT GCA GGG TCC GAG GTA TTC GCA CTG GAT ACG AC TCAGTT -3′ |

The comparative threshold cycle (Ct) was determined for each miRNA and the relative amount of each miRNA in individual samples was described as ΔCt (Ct miRNA- Ct internal control). $2^{-\Delta Ct}$ values were employed for expression level comparison of miRNAs in breast samples.
