## Supplementary figures and images for "A panel of four miRNAs (miR-190b, miR-584-5p, miR-452-5p, and miR-1306-5p) is capable of classifying luminal and non-luminal breast cancers"

### Supplementary Fig 1

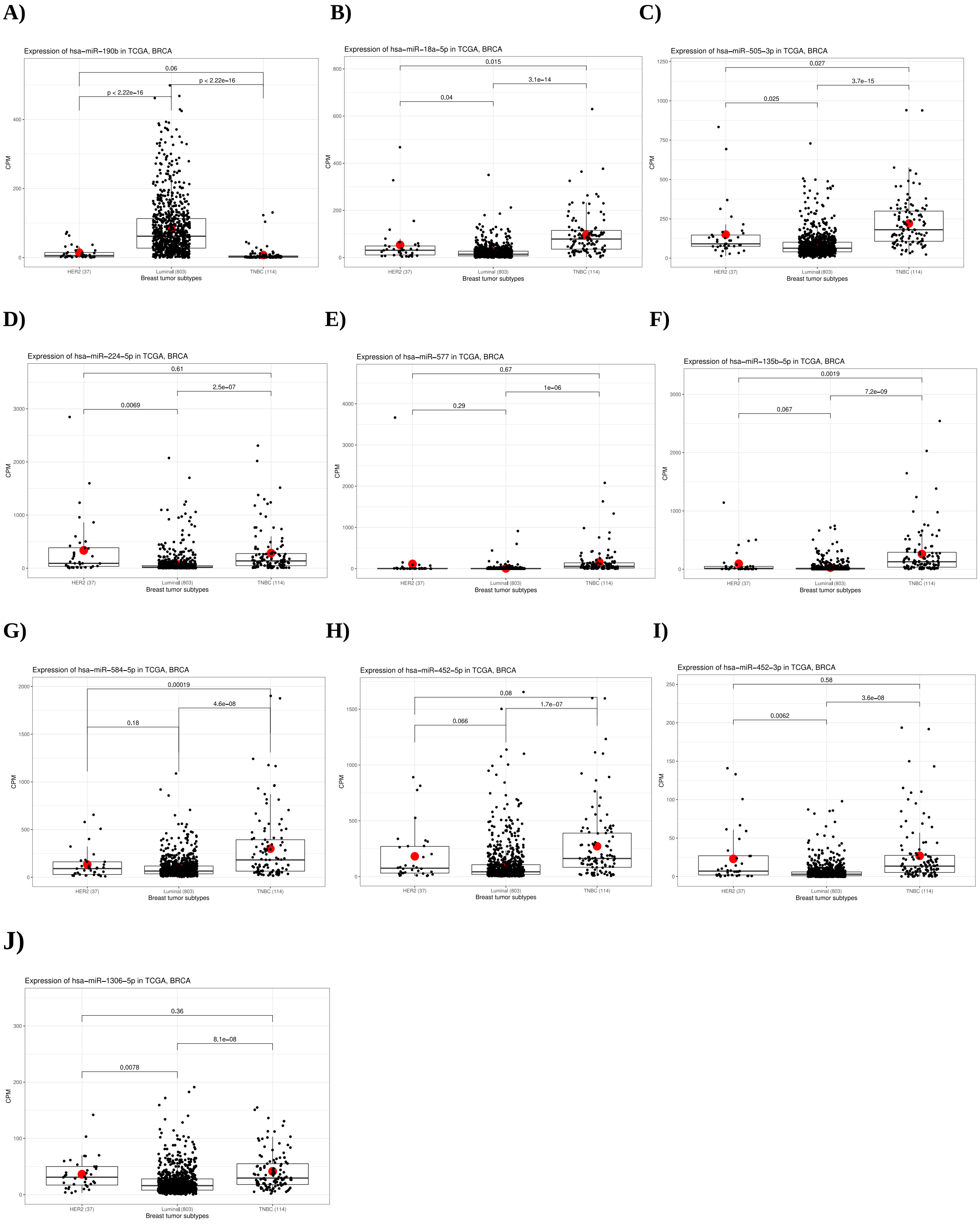

### Supplementary Fig 2

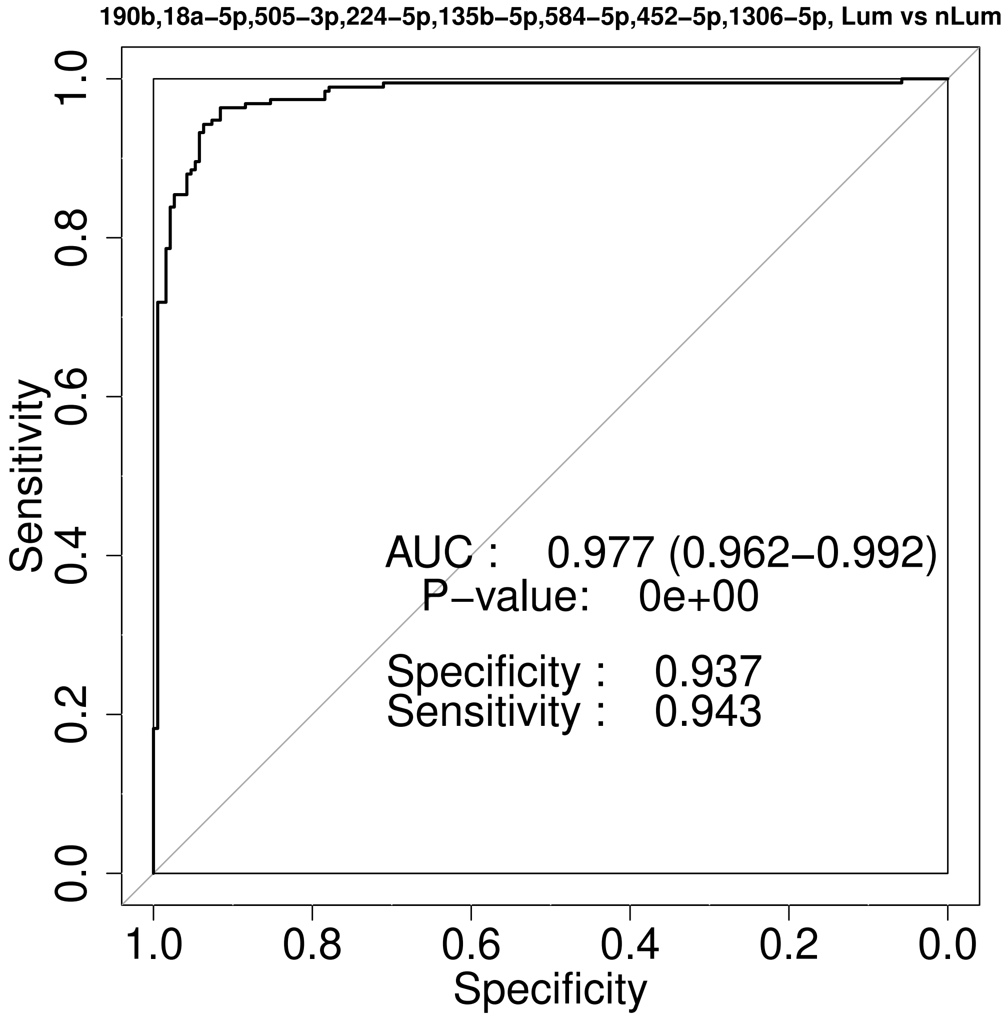

### Supplementary Fig 3

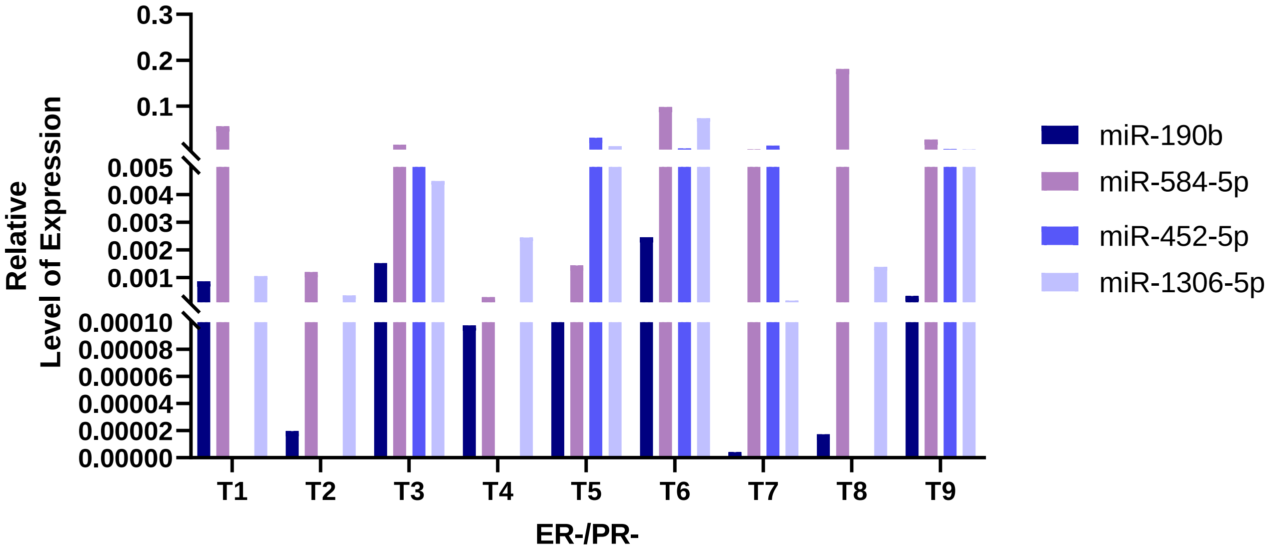

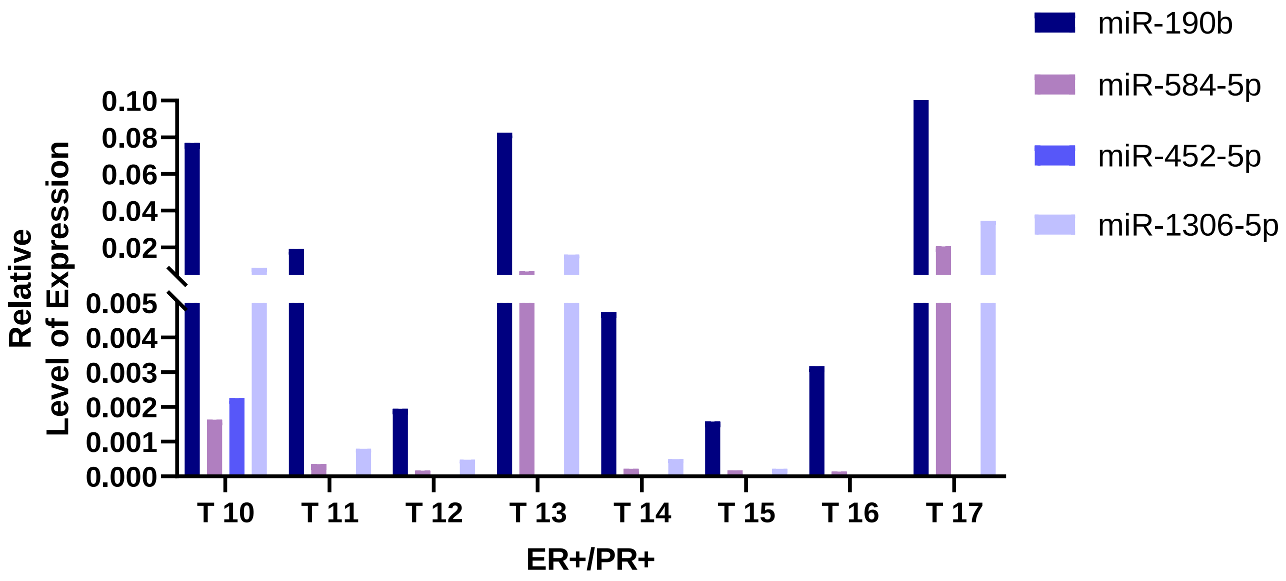
